# A Reproducible Clinical Decision-Support Suite on MIMIC-IV

**DOI:** 10.64898/2026.06.23.26356380

**Authors:** Jayakanth S. Kesan

## Abstract

This work is a direct extension and modernisation of zMed’s 2021 clinical AI/ML modelling effort, which established the company’s first intensive-care risk models on earlier critical-care data. Here we re-platform that work on the latest MIMIC-IV v3.1 release and substantially broaden it. Most published clinical-AI results are single models on a single dataset, difficult to reproduce, and rarely validated outside their training hospital. We built a broad, methodologically rigorous, reproducible clinical decision-support (CDS) suite spanning four families—intensive-care deterioration and outcomes, emergency-department triage, electrocardiographic interpretation, and clinical natural-language processing—comprising 26 models. Tabular models are gradient-boosted trees over point-in-time, leakage-safe first-24-hour features; deep models include one-dimensional convolutional networks on raw 12-lead ECG, fine-tuned clinical transformers, and an instruction-tuned large language model for discharge-summary drafting. Every model uses patient-level data splits, probability calibration, a shuffled-label leakage gate, and SHAP explanations, and is characterised by its full confusion matrix with sensitivity, specificity and predictive values. Discrimination matched or approached published benchmarks: ICU mortality AUROC 0.884, acute kidney injury 0.830, prolonged stay 0.813; emergency-department-to-ICU 0.875; cardiologist-labelled ECG diagnosis 0.909; full-note diagnostic coding 0.892. Raw-signal ECG deep learning improved myocardial-infarction detection by +0.142 AUROC over interval features. The MIMIC-trained mortality model generalised to a different multi-centre US cohort (199,133 stays) with only a 0.044 AUROC drop. We describe how each model family is incorporated into the latest version of the zMed Critical Care application and its CDS tools.

## 1 Introduction

Clinical decision support spans many tasks—predicting deterioration, triaging arrivals, reading electrocardiograms, coding notes—yet most studies tackle one task on one dataset. Three recurring weaknesses undermine trust: information leakage (features that covertly encode the outcome), absent calibration (scores that are not probabilities), and no external validation (models that work only at the hospital that trained them).

zMed first developed intensive-care risk models in 2021 as part of its clinical-AI roadmap. The present study extends that 2021 work in three ways: it re-platforms the models on the current MIMIC-IV v3.1 release [1] (which postdates the original effort); it broadens scope from ICU risk alone to emergency triage, 12-lead ECG and clinical language; and it adds the rigour the original lacked—leakage gates, calibration, SHAP explanations [11] and cross-hospital external validation. We treat the work as auditable research: every model emits a model card with its full confusion matrix, and every reported result is reproducible. Prespecified validation checks caught and corrected two latent defects—a temporal-leakage acute-kidney-injury label and a calibration-on-test-set error—before full-scale evaluation, preventing inflated performance.

## 2 Related Work and Comparison

Strong single-task models exist for each task. For ICU mortality and length-of-stay, the MIMIC-III multitask benchmark [7] reports mortality AUROC of about 0.86 on a single dataset without external validation. For acute kidney injury (AKI), Tomašev *et al*. [12] achieved AUROC of about 0.92—but on a non-public, single-health-system cohort that is not reproducible. For sepsis, interpretable ICU models [13] and the PhysioNet/CinC 2019 Challenge [14] report AUROC of 0.83–0.85. For ECG, cardiologist-level arrhythmia detection [15] used proprietary single-lead data, while 12-lead deep networks [16] and the PTB-XL benchmark [4] report macro-AUROC of about 0.93.

We do not claim to beat every specialised model on its own task. A model tuned and tested on one task and one private cohort can post a higher peak AUROC than a generalist suite. Our contribution is different and, for deployment, arguably more useful: breadth, rigour, external validation, reproducibility and translation into a live product, together (Table 1). Briefly: *breadth* (structured, waveform and text CDS in one reproducible pipeline); *leakage-safe* construction (AKI and Sepsis-3 defined as post-window onsets; every model passes a shuffled-label gate near 0.50); *calibrated and fully characterised* (Brier scores, calibration curves, and the complete confusion matrix—sensitivity, specificity, PPV and NPV—for every model); *externally validated* (the mortality model tested unchanged on a different hospital network, eICU, 199,133 stays); *reproducible* (public data and versioned code); and *deployed* (wired into a production clinical application).

**Table 1.**
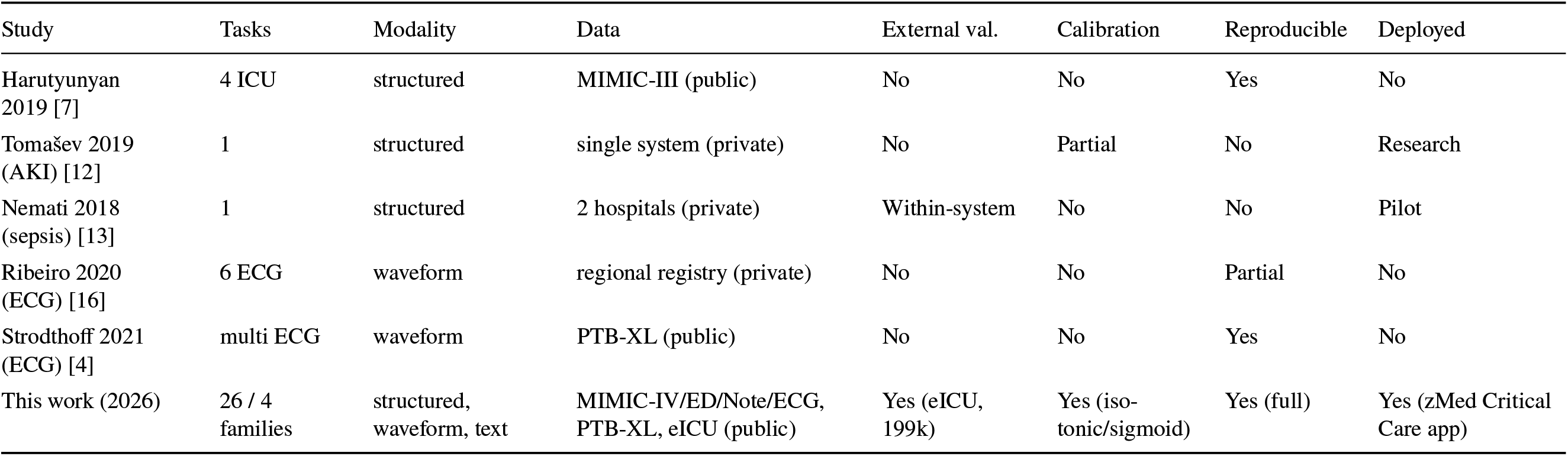
Comparison with representative prior studies.

## 3 Methods

### 3.1 Cohorts and outcomes

The ICU cohort is one row per ICU stay, anchored at admission. Outcomes: in-hospital mortality; KDIGO acute kidney injury [6]; prolonged stay (longer than 7 days); 30-day readmission; and Sepsis-3 [5] (suspected infection plus an acute SOFA rise of at least 2), following established database operationalisations of these critical-care concepts [19]. MIMIC timestamps are shifted per patient but internally consistent, so all features use relative offsets within an encounter.

### 3.2 Leakage-safe features

Tabular models observe only the first 24 hours of the ICU stay (or the triage instant, for the emergency department), summarising each vital sign and laboratory value as minimum, maximum, mean, last, and count. AKI and Sepsis-3 are detected after the 24-hour window, so inputs cannot trivially contain the label. A shuffled-label negative control gates every model: permuting labels must collapse AUROC to about 0.50; all models pass.

### 3.3 Models and evaluation

Tabular models are class-weighted gradient-boosted trees (XGBoost) [8] with isotonic/sigmoid calibration. ECG models are a five-block one-dimensional convolutional neural network on raw 12-lead signals (MIMICIV-ECG and PTB-XL). Language models comprise TF-IDF and gradient-boosted-tree baselines, fine-tuned clinical BERT [9] and Longformer-4096 [10] for diagnostic-chapter coding, and an instruction-tuned large language model [17] (zero-shot and low-rank-adapted [18]) for discharge-summary drafting scored by ROUGEL. We use patient-level 60/20/20 (tabular) or 80/20 (deep) splits so no patient appears in two folds, and report AUROC with bootstrap 95% confidence intervals, area under the precision-recall curve (AUPRC), Brier score, the confusion matrix at the Youden-optimal threshold with derived sensitivity, specificity, positive and negative predictive value (PPV, NPV), F1 and the Matthews correlation coefficient (MCC), SHAP attributions, and subgroup AUROC by sex and age. The mortality model was retrained on the 92 features common to MIMIC-IV and eICU [2] and evaluated, unchanged, on the entire eICU cohort.

## 4 Results

### 4.1 Discrimination across the suite

Discrimination is summarised in Figures 1 and 2 and detailed in Tables 2 and 3. ICU mortality reached 0.884 (95% CI 0.876–0.891), AKI 0.830, prolonged length-of-stay 0.813, readmission 0.616, and incident Sepsis-3 [5] 0.794. Emergency-department-to-ICU reached 0.875 and to-admission 0.802 from triage data alone. SHAP attributions are clinically coherent (mortality driven by age, blood urea nitrogen, lactate and mean arterial pressure; AKI by the creatinine trajectory).

**Table 2.** Catalogue of CDS models: clinical use, inputs, method and headline result.

| Model | Clinical usage | Input clinical parameters | Method | Result |
| --- | --- | --- | --- | --- |
| ICU mortality | risk flag, escalation | first-24h vitals + labs<br>(min/max/mean/last/count), age, sex,<br>emergency | XGBoost<br>(calibrated) | AUROC 0.884 |
| ICU AKI<br>(post-24h) | early kidney-injury<br>warning | first-24h creatinine + full panel | XGBoost | 0.830 |
| ICU prolonged stay | capacity / step-down | first-24h vitals + labs | XGBoost | 0.813 |
| ICU 30-day<br>readmission | discharge planning | first-24h vitals + labs | XGBoost | 0.616 |
| ICU Sepsis-3<br>(incident) | sepsis surveillance | first-24h SOFA inputs +<br>suspicion-of-infection | XGBoost + SOFA | 0.794 |
| ED to admission | disposition at triage | triage vitals, acuity, arrival mode, age, sex | XGBoost | 0.802 |
| ED to ICU | ICU-need flag | triage vitals + acuity | XGBoost | 0.875 |
| ED high acuity | under-triage safety<br>net | triage vitals (acuity excluded) | XGBoost | 0.760 |
| Diagnostic coding<br>(3 chapters) | auto-coding | discharge-note text | TF-IDF +<br>XGBoost | 0.94 / 0.83 /<br>0.85 |
| Coding<br>(transformer) | richer coding | discharge note (512 tokens) | clinical BERT [9] | macro 0.885 |
| Coding<br>(long-context) | whole-note coding | discharge note (4096 tokens) | Longformer-4096 | macro 0.892 |
| Discharge-summary draft | draft course for edit | note text before course | LLM + LoRA | ROUGE-L<br>0.070 to 0.144 |
| ECG 12-condition | abnormality screen | device ECG intervals/axes | XGBoost | 0.74–1.00 |
| ECG raw-signal<br>(12) | signal-level screen | raw 12-lead waveform | 1-D CNN | macro 0.926 |
| ECG diagnosis (5) | diagnostic triage | raw 12-lead (PTB-XL) | 1-D CNN | macro 0.909 |
| Mortality (external) | cross-hospital<br>transfer | 92 shared vitals + labs | XGBoost | 0.880 to 0.836 |

**Table 3.** Full operating-point performance for the twelve re-runnable tabular models (Youden-optimal threshold, held-out test sets). The AUROC column is the single-split re-run and differs from the calibrated-pipeline headline (Table 2) by at most 0.01.

| Model | Test $n$ | Prev. | AUROC | AUPRC | Sens. | Spec. | PPV | NPV | F1 | MCC |
| --- | --- | --- | --- | --- | --- | --- | --- | --- | --- | --- |
| ICU mortality | 28,365 | 0.121 | 0.883 | 0.604 | 0.79 | 0.80 | 0.35 | 0.97 | 0.49 | 0.43 |
| ICU AKI (KDIGO) [6] | 17,177 | 0.268 | 0.831 | 0.692 | 0.72 | 0.78 | 0.55 | 0.88 | 0.62 | 0.47 |
| ICU prolonged stay | 28,347 | 0.120 | 0.809 | 0.384 | 0.73 | 0.74 | 0.27 | 0.95 | 0.40 | 0.33 |
| ICU 30-day readmission | 28,365 | 0.184 | 0.612 | 0.262 | 0.54 | 0.62 | 0.24 | 0.86 | 0.34 | 0.13 |
| ICU Sepsis-3 (incident) | 8,611 | 0.052 | 0.796 | 0.180 | 0.91 | 0.55 | 0.10 | 0.99 | 0.18 | 0.21 |
| ED to admission | 128,549 | 0.373 | 0.802 | 0.709 | 0.72 | 0.73 | 0.61 | 0.81 | 0.66 | 0.44 |
| ED to ICU | 128,549 | 0.075 | 0.874 | 0.422 | 0.80 | 0.78 | 0.23 | 0.98 | 0.35 | 0.35 |
| ED high acuity | 125,143 | 0.391 | 0.756 | 0.697 | 0.65 | 0.72 | 0.60 | 0.76 | 0.62 | 0.37 |
| Coding: circulatory | 5,895 | 0.719 | 0.947 | 0.979 | 0.86 | 0.91 | 0.96 | 0.72 | 0.91 | 0.72 |
| Coding: respiratory | 5,895 | 0.326 | 0.837 | 0.763 | 0.73 | 0.81 | 0.66 | 0.86 | 0.69 | 0.53 |
| Coding: infectious | 5,895 | 0.231 | 0.837 | 0.636 | 0.74 | 0.78 | 0.50 | 0.91 | 0.60 | 0.46 |
| Mortality (external, eICU) | 199,133 | 0.092 | 0.838 | 0.492 | 0.77 | 0.73 | 0.22 | 0.97 | 0.35 | 0.31 |

**Figure 1.**
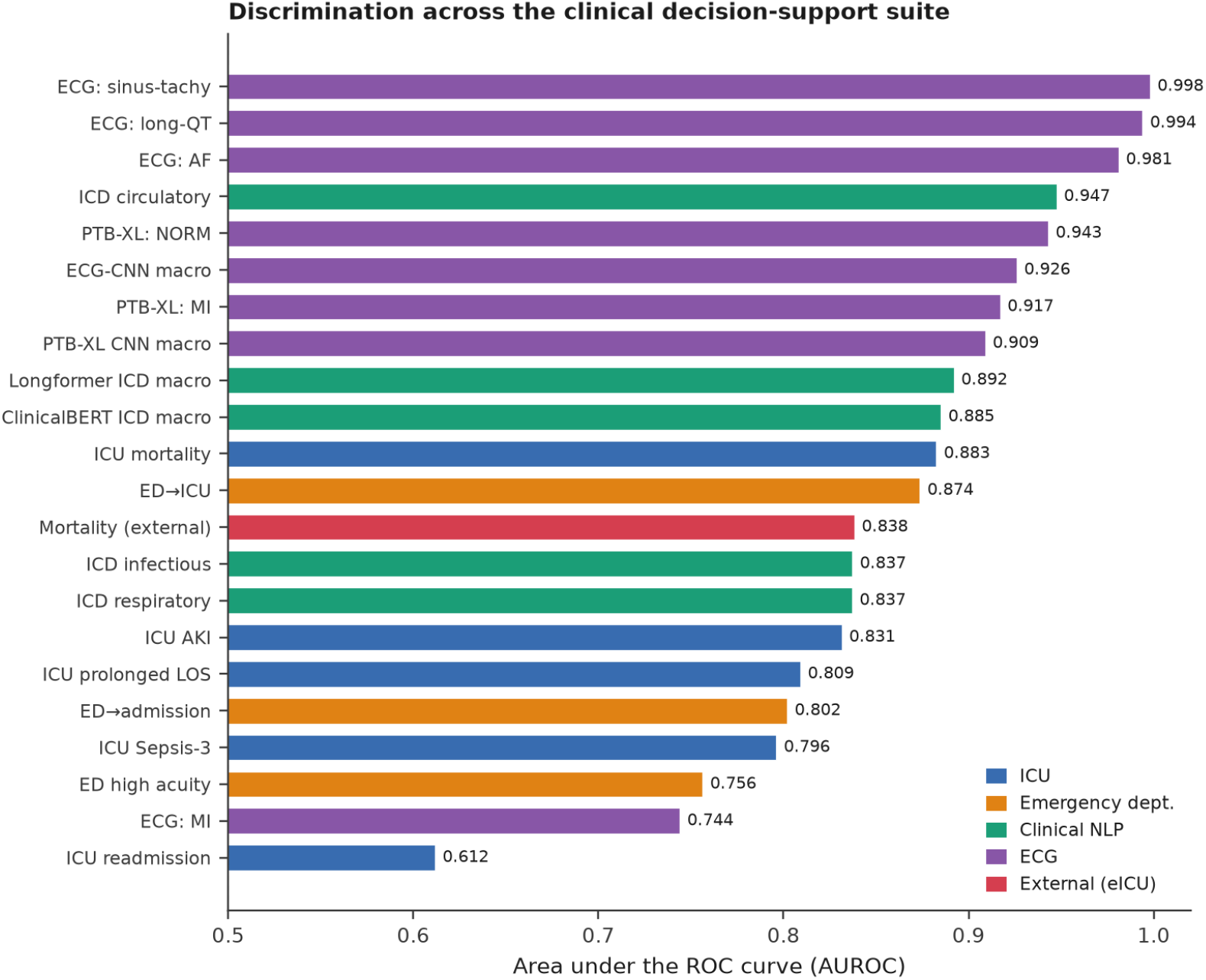
Discrimination (area under the ROC curve) across the evaluated decision-support model heads, coloured by clinical family. The dashed line marks chance (0.50). Bars for the re-runnable tabular models are single-split values and differ from the calibrated-pipeline headline (Table 2) by at most 0.01—a few thousandths for most models—owing to stochastic patient-level splitting.

**Figure 2.**
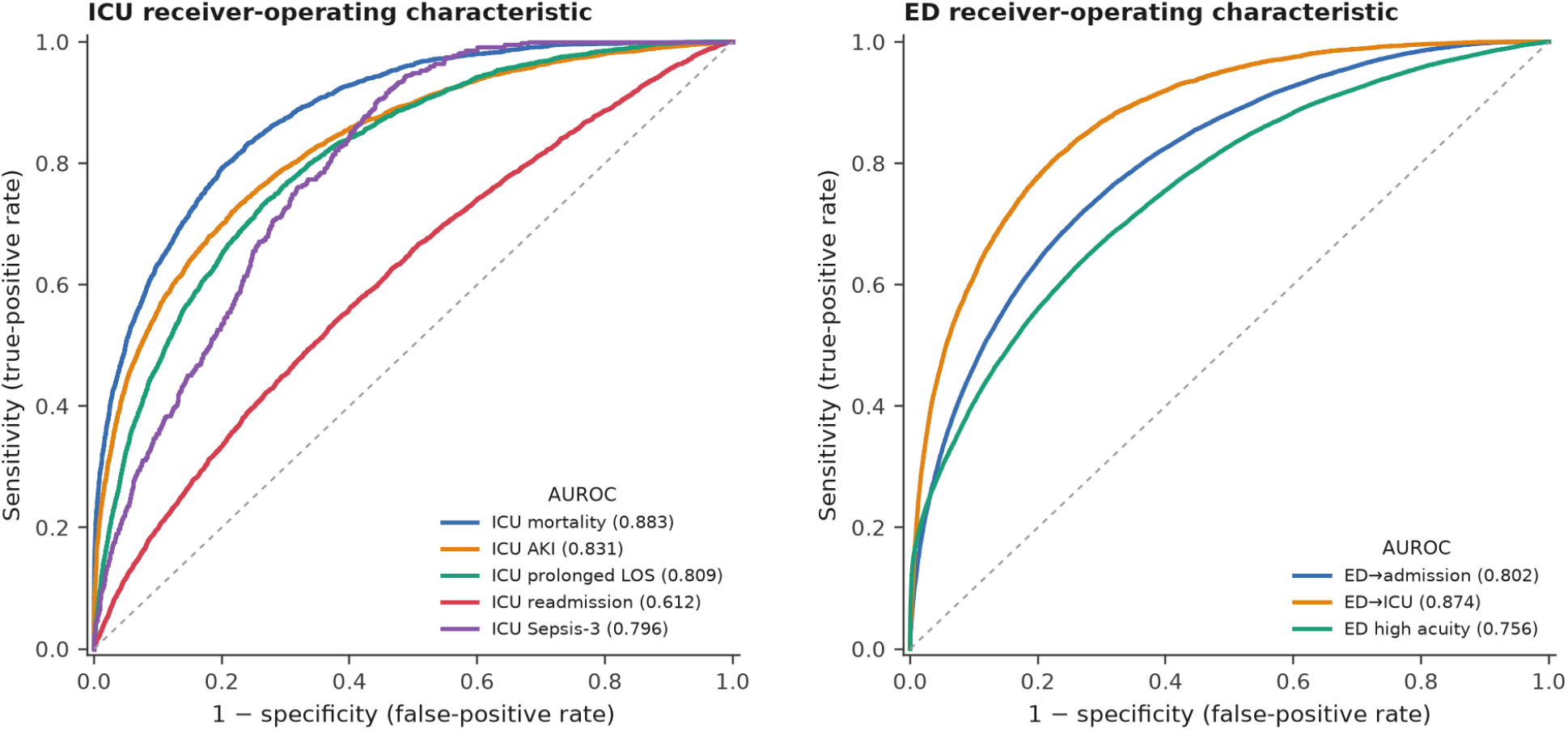
Receiver-operating-characteristic curves for the intensive-care (left) and emergency-department (right) decision-support models on held-out test sets. Each curve plots sensitivity against 1–specificity; the diagonal marks chance (AUROC of 0.50). AUROC values appear in the legends.

### 4.2 Operating-point behaviour

Beyond ranking, a deployable model must be characterised at a decision threshold. Table 3 and Figures 3 and 4 give the full confusion matrix at the Youden-optimal operating point for the twelve re-runnable tabular models—true and false positives and negatives, sensitivity, specificity, positive and negative predictive value, F1 and the Matthews correlation coefficient—together with each test cohort’s size and outcome prevalence. The pattern is the expected one for screening CDS on imbalanced outcomes: high sensitivity and very high negative predictive value (at or above 0.95 for the rare ICU and ED-to-ICU outcomes), with positive predictive value bounded by low prevalence. ICU mortality (prevalence 0.12) achieves sensitivity 0.79, specificity 0.80 and NPV 0.97; incident Sepsis-3 (prevalence 0.05) reaches sensitivity 0.91 and NPV 0.99. Where prevalence is high, precision rises accordingly (diagnostic coding of circulatory disease: PPV 0.96, F1 0.91).

**Figure 3.**
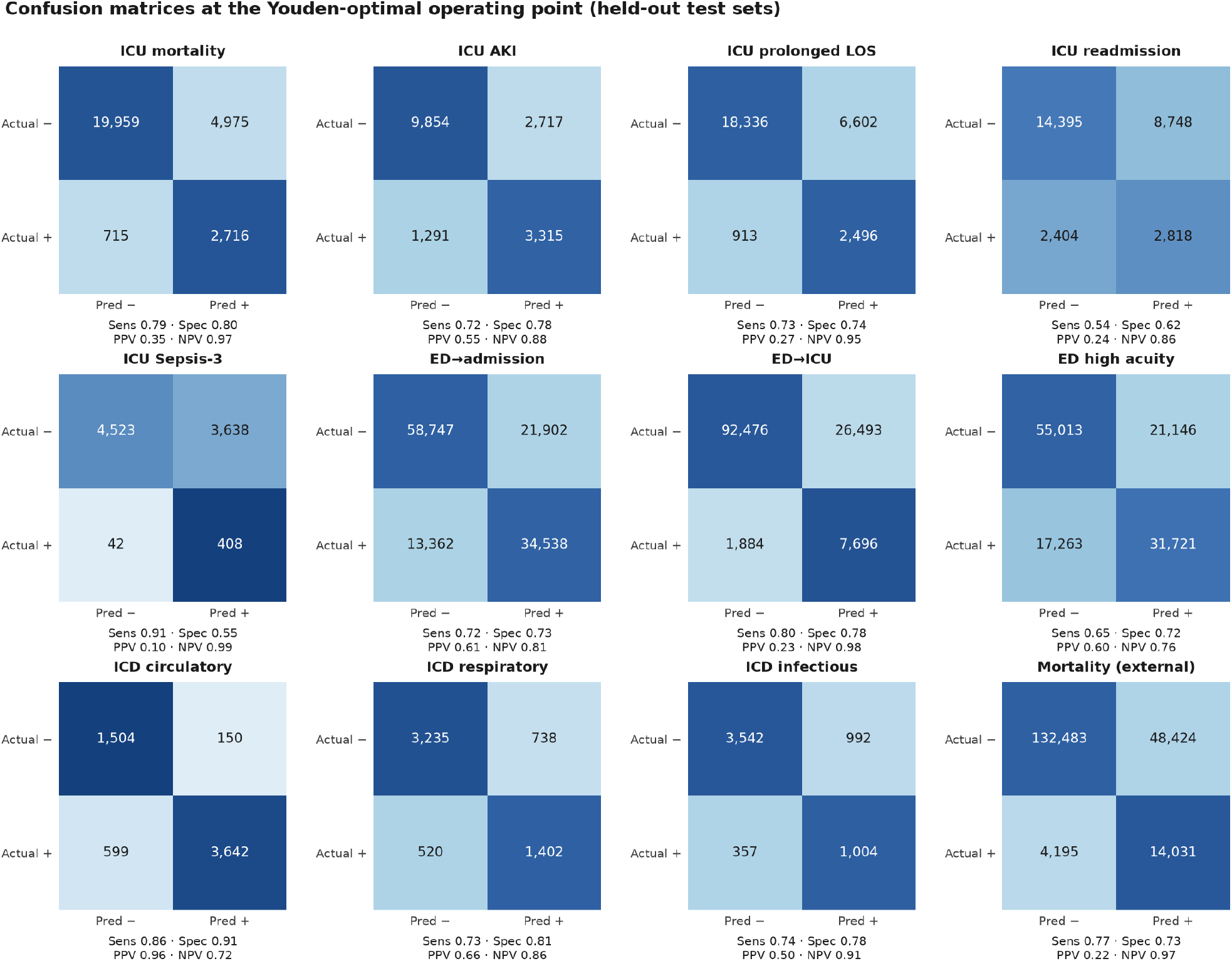
Confusion matrices at the Youden-optimal operating point on held-out test sets. Cell colour is row-normalised (per actual class); annotations are raw counts. Each panel reports sensitivity, specificity, PPV and NPV. Rows: actual class; columns: predicted class.

**Figure 4.**
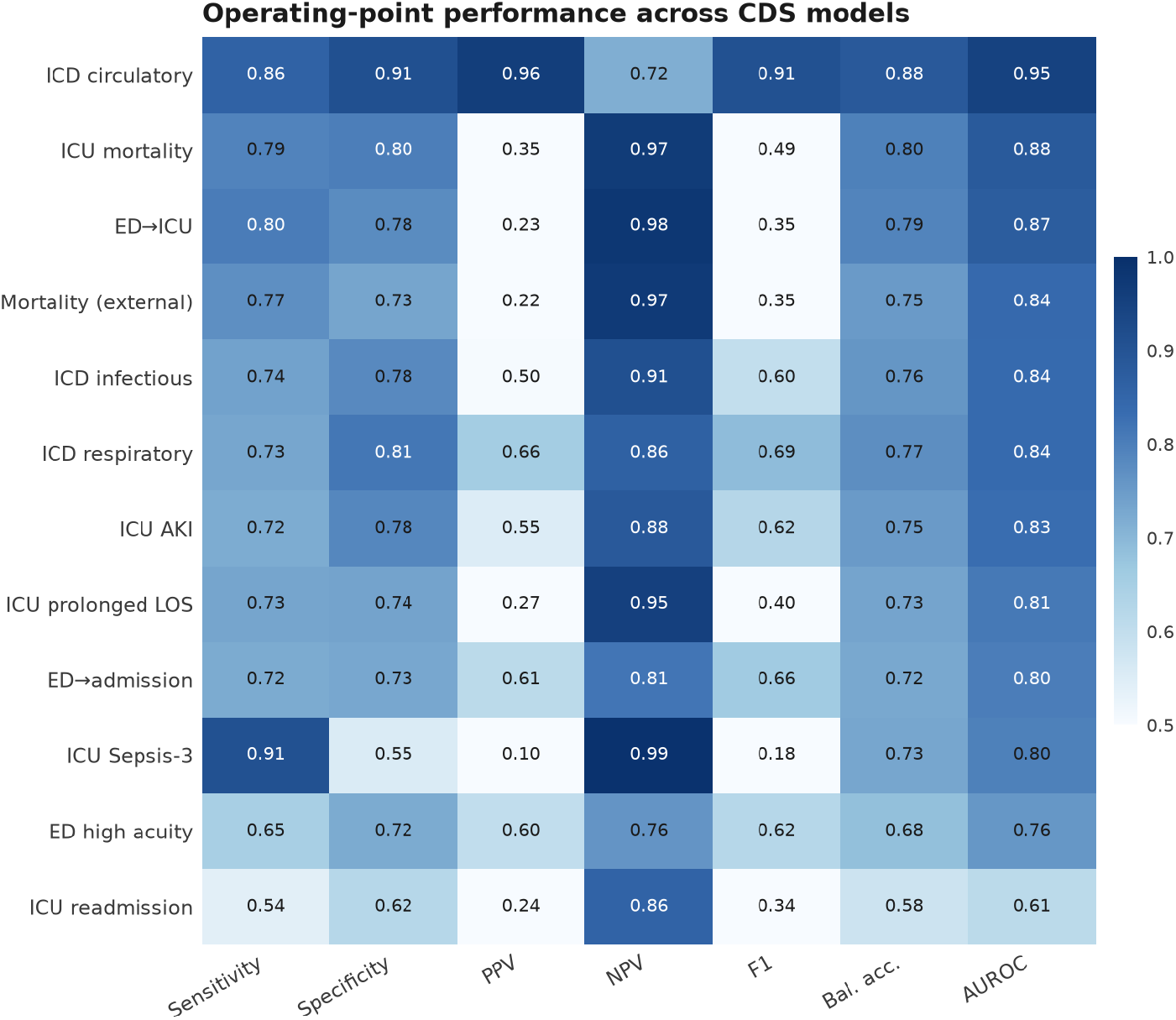
Operating-point performance (sensitivity, specificity, PPV, NPV, F1, balanced accuracy and AUROC) across all twelve tabular models, ordered by AUROC. The deep-learning models (ECG CNN, PTB-XL, transformer coding) contribute discrimination only; their per-case predictions were not retained for this operating-point analysis.

### 4.3 Waveform, language and transfer

*ECG*. An interval and axis model over 800,000 ECGs spanned AUROC 0.74–1.00 across twelve conditions; a raw-signal CNN reached macro-AUROC 0.926 and improved myocardial-infarction detection to 0.886 (+0.142), capturing ST/T morphology that interval features cannot see. On cardiologist-labelled PTB-XL [3] the same architecture achieved macro-AUROC 0.909. *Language*. TF-IDF diagnostic-chapter coding reached 0.94/0.83/0.85; a Longformer-4096 [10] (macro 0.892) added long-range context, helping respiratory and infectious categories. Discharge-summary drafting improved from ROUGE-L 0.070 (zero-shot) to 0.144 with low-rank fine-tuning [18]. *External validation*. The mortality model transferred to eICU [2] (199,133 stays) with internal AUROC 0.880 to external 0.836 (gap 0.044), strong evidence it is not overfit to one hospital.

## 5 Discussion

Four findings stand out. First, raw-waveform deep learning earns its keep precisely where engineered features fail (the +0.142 AUROC for myocardial infarction). Second, the suite generalises—a 0.044 external-validation drop is the number that speaks to deployability, and the consistently high negative predictive values (Table 3) make these models well-suited to their primary clinical role of safely ruling out low-risk patients. Third, long-context language models help selectively, where evidence is dispersed through a note. Fourth, fine-tuning, not scale, is the lever for grounded summarisation. Methodologically, the project ar-gues for treating clinical AI as auditable research, with calibration, leakage gates, external validation and full confusion-matrix reporting as defaults rather than extras.

The work has clear limitations. MIMIC training data are single-centre; despite the eICU transfer, prospective multi-site validation is required before clinical use. The 800,000-ECG metadata labels are device-generated, mitigated by the cardiologist-labelled PTB-XL analysis. Sepsis-3/SOFA and KDIGO are operationalised approximations of the reference concepts. ROUGE-L is a weak proxy for summary quality. The detailed operating-point analysis (Table 3) covers the twelve re-runnable tabular models; the deep-learning models report discrimination only, because their per-case predictions were not retained for operating-point analysis. None of these models is a regulated medical device; all results are research artefacts on retrospective, de-identified data.

## 6 Translation Into Deployed Clinical Care

The models are designed for incorporation into the latest version of the zMed Critical Care application and its CDS tools. The application already operates a decision-support layer that combines rule-based and statistical alerting, surfaced to clinicians at the bedside. This work upgrades that layer from heuristics to validated, versioned, calibrated machine-learning models with an auditable performance pedigree.

### Validated model registry

Each ICU and ED model is registered with its measured discrimination, sensitivity, specificity and cohort size (Table 3), so every score is traceable to a documented evaluation, and each prediction is logged for audit. *Real-time scoring*. The same first-24-hour (or triage-instant) feature contract used here is computed online from the data already flowing through the application; risk scores are written back to the patient record and raised as severity-ranked, click-to-patient alerts. *Waveform and ECG*. The raw-signal ECG network complements the application’s existing real-time waveform monitoring, adding diagnostic-grade screening alongside rhythm alarms. *Notes and language*. The coding and discharge-summary models extend the application’s clinical-documentation tooling and are available conversationally through its built-in clinical AI assistant. *Validation environment*. A de-identified MIMIC cohort is maintained inside the application as an isolated, permanent regression and demonstration environment— de-identified data only, never mixed with real patient records.

### Regulatory positioning: human-in-the-loop and Non-Device CDS

The deployment architecture is built on a human-in-the-loop principle: the models never act autonomously; every output is a recommendation to a licensed clinician, who makes the decision. Each recommendation is displayed with the basis for it—the underlying clinical inputs, the calibrated probability, the operating point with its sensitivity and specificity (Table 3), and the SHAP attribution of the principal drivers—so the clinician can review the reasoning and accept or override it. This design is intended to support a Non-Device Clinical Decision Support determination under section 520(o)(1)(E) of the U.S. Federal Food, Drug, and Cosmetic Act—added by the 21st Century Cures Act—as interpreted by the FDA’s 2022 final guidance on Clinical Decision Support Software [20,21]. We are deliberately conservative about which functions this covers (Table 4): documentation and reference aids are the strongest Non-Device CDS candidates, whereas patient-specific risk scores for serious or time-critical conditions and any function that analyses a physiological signal directly are treated as Software as a Medical Device (SaMD) rather than asserted as exempt. This classification is jurisdiction-specific: the section 520(o) carve-out is United States only, whereas the EU Medical Device Regulation (Rule 11) and India’s CDSCO regime generally classify such decision-support software as a regulated medical device with no equivalent exemption. zMed is in the process of acquiring the necessary certifications and clearances for these models in the relevant jurisdictions. Until those are in place, all models described here remain research artefacts pending prospective validation and regulatory authorisation, and are not offered for autonomous clinical use.

**Table 4.** Regulatory classification of the deployed CDS functions.

| Deployed function | Likely regulatory class | Pathway / status |
| --- | --- | --- |
| Diagnostic-coding suggestions | Non-Device CDS candidate | Documentation aid; exempt where the four criteria are met |
| Discharge-summary drafting | Non-Device CDS candidate | Clinician-edited draft; exempt where criteria met |
| ICU mortality / deterioration risk | Software as a Medical Device | Certification / clearance in progress |
| Sepsis-3 early warning | Software as a Medical Device | Certification / clearance in progress |
| AKI prediction | Software as a Medical Device | Certification / clearance in progress |
| ED triage (admission / ICU) | Software as a Medical Device | Certification / clearance in progress |
| Raw 12-lead ECG analysis | Software as a Medical Device (signal analysis) | Device pathway (e.g., 510(k) / CE / CDSCO) |

## Data Availability

All data produced in the present work are contained in the manuscript and references

## Declarations

### Ethics approval

This study is a secondary analysis of fully de-identified, publicly available datasets (MIMICIV, eICU-CRD, PTB-XL) accessed under their respective data use agreements. It does not constitute humansubjects research and was exempt from institutional review board approval; no additional patient consent was required.

### Data availability

MIMIC-IV and eICU-CRD are available via PhysioNet to credentialed users under a signed data use agreement; PTB-XL is openly available on PhysioNet. No patient-level data are shared in this manuscript; only aggregate results are reported.

### Code availability

The analysis pipeline (feature derivation, models, evaluation, figures and the operating-point tables) re-runs from a documented command set and is available from the author on reasonable request.

### Author contributions

J.S.K. conceived the study, implemented the pipeline, performed the analysis, and wrote the manuscript.

### Reporting

Prediction-model results are reported in line with the TRIPOD+AI recommendations.

